# ‘Whose quality of life is it anyway?’ – Evaluation of quality of life tools for children with complex needs accessing specialist leisure provision

**DOI:** 10.1101/2023.11.15.23298564

**Authors:** Fiona Astill, Bethan Collins, Nicole McGrath, Alison Kemp, Lisa Hurt, Sabine Maguire

## Abstract

Studies of quality of life (QoL) routinely exclude children with complex needs. These children struggle to access leisure activities, particularly those with severe communication needs or challenging behaviour. Sparkle provides specialised leisure services to children and young people (0-17 years) with complex needs in South Wales, UK. We aimed to evaluate previously validated tools to measure QoL with this population.

Three tools were assessed over a 6-year period – PedsQL, KINDL^R^ and QI-Disability. PedsQL (41) and KINDL^R^ (10) were attempted by the children attending the clubs (5-17 years old), and QI-Disability by caregivers (96). The majority of child participants had a neurodevelopmental diagnosis, a proportion of whom were non-verbal.

Neither KINDL^R^ nor PedsQL were appropriate for the population, with children unable to understand the questions and answers. The QI-Disability scores showed a statistically significant improvement in parents’ estimate of their child’s positive emotions, but results were severely limited by drop off.

Existing validated QoL tools cannot be meaningfully used by children with complex needs. While the caregiver tool showed some benefit of specialist leisure provision, it is recognised that caregivers may perceive a child’s QoL differently to the child themselves, and caregivers clearly found repeat measurements onerous.

## Introduction

Play is an essential right for all children (UNICEF, 1992) and provides an opportunity to develop vital social and problem-solving skills (Lewis, 1993). However, children with complex needs often have limited access to play, and may not experience the same benefit from toys or routine play activities as their non-disabled peers, thus requiring specialist provision or adjustments to see the most benefit (Dahan-Oliel et al., 2012). A recent systematic review exploring the meaning of play for children with physical disabilities highlighted that these children experience play differently, and recognise that they expect to need help to access play activities (Graham et al., 2018). However, there are no reports in the literature of supported play opportunities for children with disability or cognitive impairment, where they are allowed to choose and influence which play activities they engage in.

Sparkle (South Wales) is the partnered charity of Serennu, Nevill Hall and Caerphilly Children’s Centres; Sparkle supports children and young people (aged 0-17 years) with a range of disabilities and developmental difficulties, and their families, in part by delivering specialised leisure activities. The aim of this is to provide these children and young people with the same leisure opportunities as other children, empowering them by enabling them to choose the play activities, and supporting them to develop social skills and independence, and improve their wellbeing, including the opportunity to develop meaningful friendships.

The World Health Organisation (WHO) defines Quality of Life (QoL) as the perception of an individual’s life *“in the context of the culture and value systems in which they live and in relation to their goals, expectations, standards and concerns”* (WHO, 2022). For individuals with a chronic health condition or an underlying disorder that can impact on their ability to interact with society, or society’s perception of them, this is an additional influencing factor on their quality of life; Health-related Quality of Life (HRQoL) is a better descriptive term for this influence. Leisure provision for children with complex needs may be ‘integrated’ where the children with complex needs participate in activities with neurotypical or non-disabled children, although activities and pace tend to be determined by the majority, i.e., able bodied/ neurotypical. Some specialist provision has also been developed, tailored to the needs of the children accessing the service. In order to determine what model of leisure provision is most effective for children with complex needs, it is essential to evaluate the impact of such provision on the children’s QoL, preferably utilising a validated tool.

Unfortunately, there is a paucity of tools suitable for measuring QoL of children and young people with disabilities, particularly those who have a cognitive or communicative disability. There is even less research relating to the value of play for children with severe emotional and/ or cognitive impairments, particularly those with significant communication difficulties (Whitebread, 2012). Indeed, it is notable that many studies assessing children’s QoL explicitly *exclude* children with significant disability or cognitive impairment or those with Autistic Spectrum Disorder or Attention Deficit Hyperactivity Disorder (Kramer et al., 2021; Longo et al., 2017; Shikako-Thomas et al., 2012; Davies et al., 2017; Dalgaard et al., 2022). Some reasons for this exclusion included children’s communication difficulties, or severe cognitive or intellectual impairment, meaning they cannot meaningfully complete self-report measures; however some authors do not explain their rationale for excluding these children, thus it is unclear if it is simply too difficult, or that the QoL of these children is somehow less important. To fill these gaps, we report on our experience of testing the use of three validated QoL tools - PedsQL, KINDL^R^ and QI-Disability - to determine the impact of a specialist, child-directed leisure provision for children with complex needs.

## Materials and Methods

Sparkle’s specialist leisure provision consists of weekly clubs, for those aged 0 to 17 years, with each session lasting 1-1.5 hours for children less than 12 years, and 2 hours for those aged >12 years. This evaluation relates to children aged 5-17 years. The clubs consist of indoor and outdoor play activities (full details in Figures 1 and 2). The aims and objectives for each club are detailed in Appendix 1. All clubs benefit from ‘enrichment activities’, including animal experiences, sports, music and dance etc. Approximately half of the children who attend require 1:1 or 2:1 support due to their significant communication or behavioural needs. The remainder receive support from trained Leisure Support Workers in small groups of up to 4 children per member of staff.

**Figure 1.**
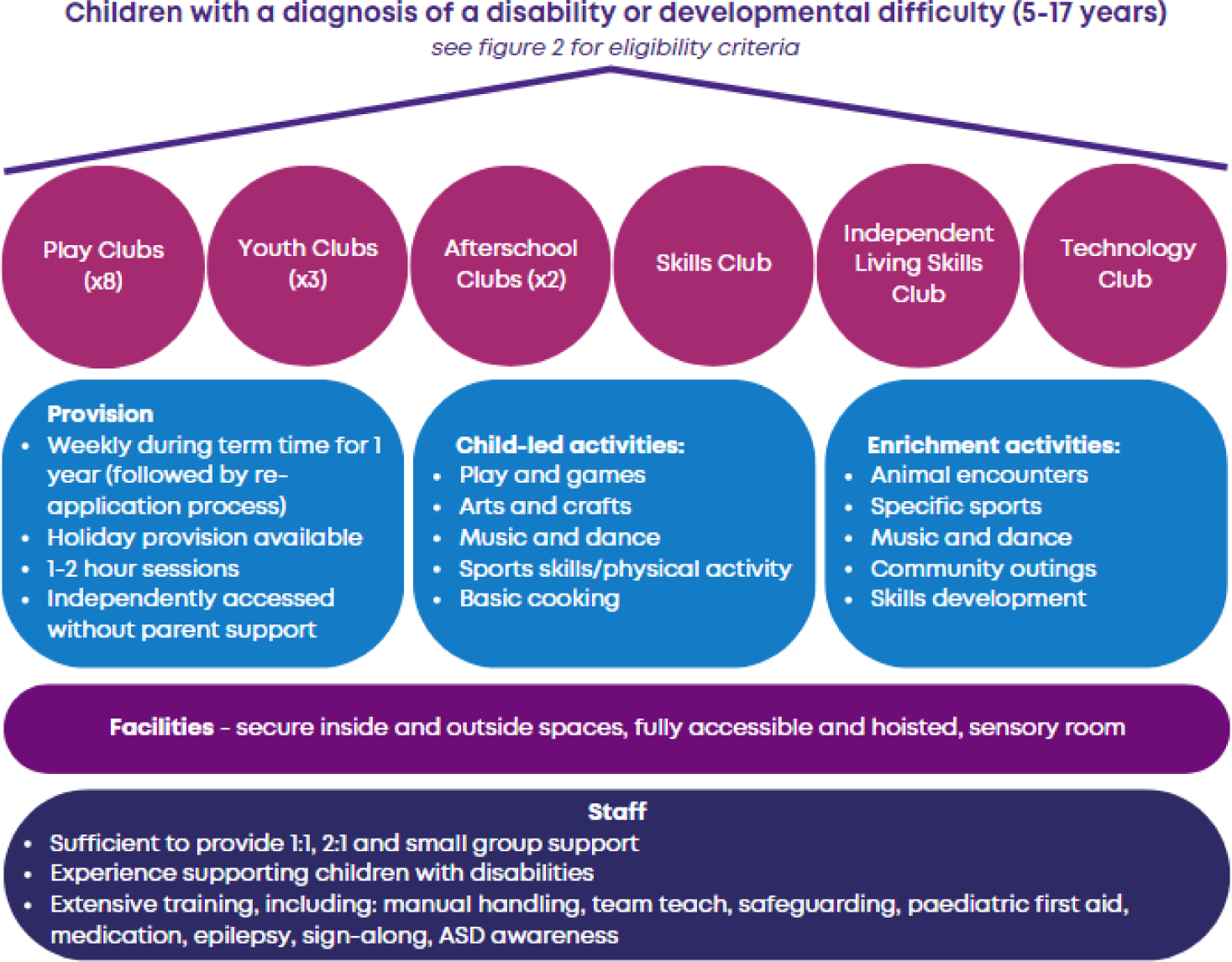
Specialist leisure provision for children aged 5-17 years with complex disabilities, provided by Sparkle (South Wales).

**Figure 2.**
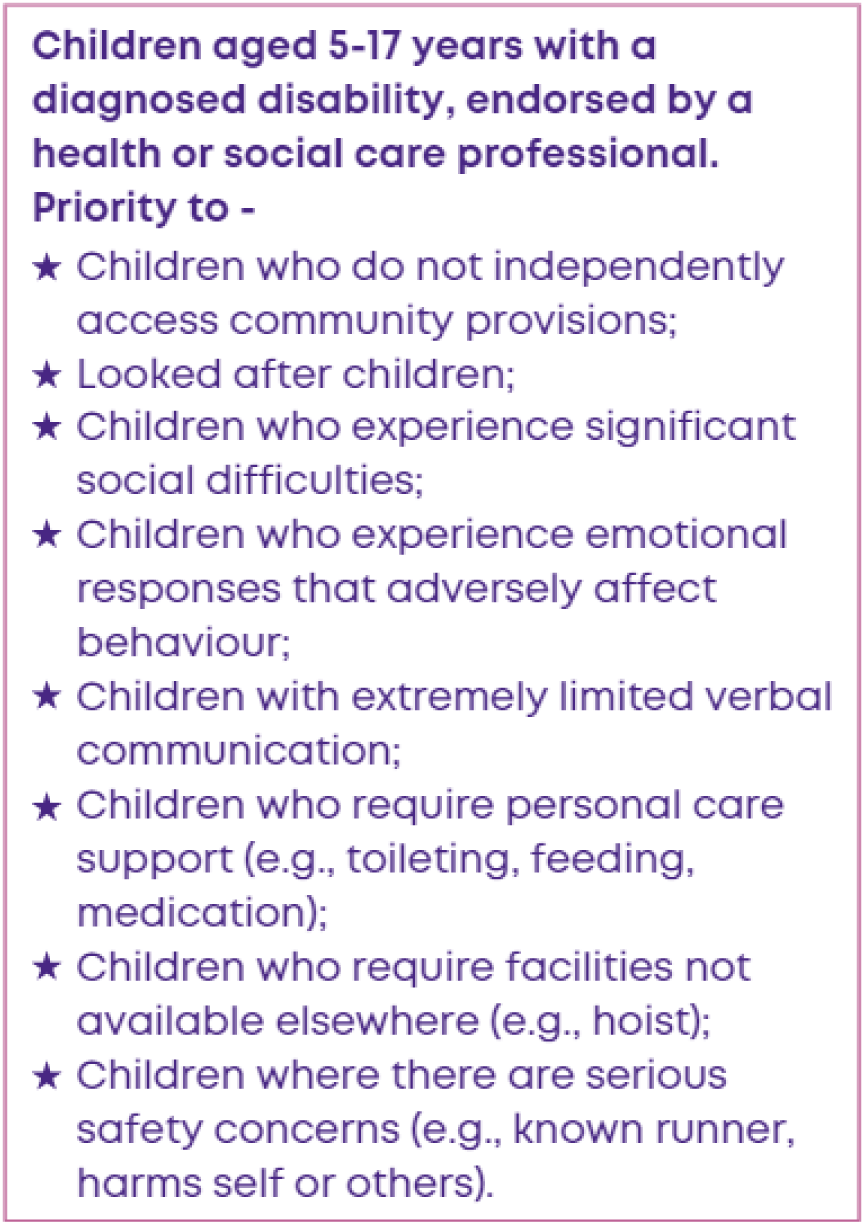
Eligibility criteria for Sparkle (South Wales)’s specialist leisure provision for children aged 5-17 years with complex disabilities.

Parents and carers (participants) of all new referrals to Sparkle’s specialist leisure activities were invited to take part in evaluations. The diagnoses of children attending the activities included Autistic Spectrum Disorder, Down’s Syndrome, Cerebral Palsy, Attention Deficit Hyperactivity Disorder, with a number of children having more than one diagnosis. It was intended that all tools would be administered prior to the child commencing their club (baseline) and again at 6 and 12 months following the child accessing a weekly club. Families were provided with a Participant Information Sheet and consent was given for participation.

Ethical approval was granted through the Research and Development Board of Aneurin Bevan Health Board; approval for the QI-Disability project was granted on 30^th^ June 2020 (SA/1146/20).

To measure the effect of specialist leisure activities delivered by Sparkle on quality of life for children and young people with complex needs, three published validated measures of quality of life for children with disabilities were evaluated sequentially over a 6 year period: Quality of Life Inventory-Disability (QI-Disability) (Telethon Kids Institute, 2022), KINDL^R^ and PEDSQL (Table 1).

**Table 1.** Quality of Life tools developed to assess Health Related Quality of Life among children and young people with complex needs, which were trialled with a group of children with significant cognitive/ communication impairments accessing a specialist leisure provision.

| *Name* | *Reference* | *Domains* | *Target group* | *Respondents* | *Age (years)* |
| --- | --- | --- | --- | --- | --- |
| PedsQL | PedsQL™ 4.0; Varni et al., (2001) | Physical Functioning<br>Emotional Functioning<br>Social Functioning<br>School Functioning | General population (child self-report and parent proxy) | Children and young people | 5-7<br>8-12<br>13-18 |
| KINDL<sup>R</sup> | Ravens-Sieberer, U., & Bullinger, M. (1998). Assessing health-related quality of life in chronically ill children with the German KINDL: First psychometric and content analytical results. Quality of Life Research, | Physical wellbeing<br>Emotional wellbeing<br>Self-esteem<br>Family<br>Friends<br>School Functioning | Disabled children and young people in mainstream education | Children and young people | 4-6<br>7-13<br>14-18 |
| QI-Disability | Telethon Kids Institute. *QI-Disability*. https://www.telethonkids.org.au/our-research/brain-and-behaviour/disability/child-disability/qi-disability/ | Positive emotions<br>Negative emotions<br>Independence<br>Physical health<br>Social Interactions<br>Leisure and the outdoors | Children and young people with neuro-developmental disorders, Retts and Down's syndrome | Caregivers | 5-18 |

The *PedsQL* is comprised of four domains, with a total of 23 items, and responses captured using a Likert scale, ranging from ‘almost always’ to ‘never’. Self-report *child* versions can be used alongside *parent* proxy versions, as a way of assessing the parent/carer’s perceptions of the child’s health-related QoL. Questions for the different age groups differed only in using developmentally appropriate language. This was tested on 9 children and 32 caregivers between 2017-2019,

The *KINDL^R^* (Ravens-Sieberer & Bullinger, 1998) is a generic instrument for assessing HRQOL in *children*, with 24 items covering six domains, reduced to 12 items for younger children (4-6 years), and three response options for each question from ‘never’ to ‘very often’. A parent/carer version, and disease specific (e.g., asthma) versions have also been developed. This was tested on 10 children between 4 and 17 years of age between October and November 2019.

The *QI-Disability* is a quality of life measure for children and young people between the ages of 5 and 18 years who have an intellectual disability. It was designed in partnership with a number of families and carers representing a wide range of intellectual disabilities as a *parent-reported* questionnaire, with the aim to evaluate the children’s and young peoples’ health and wellbeing via caregiver report. The QI-Disability matrix can have a maximum possible score of 600 across 6 domains, with a higher score indicating a higher QoL (Downs et al., 2019). The six QoL domains are assessed through 32 questions, with five responses from ‘never’ to ‘very often’. This tool was given to 96 caregivers to complete between November 2020 and September 2022.

It should be noted that during the period March 2020-December 2022, Sparkle leisure activities were restricted by the mandatory regulations relating to the COVID 19 Pandemic, as set out by the Welsh Government UK, with strict limits on numbers attending, intensive hygiene measures and social distancing of 1.5-2.0 metres which could only be achieved for these children by employing additional staff members. All of these measures, reduced the availability of spaces for children to attend the specialist provision (see participant flowchart, Figure 3).

**Figure 3.**
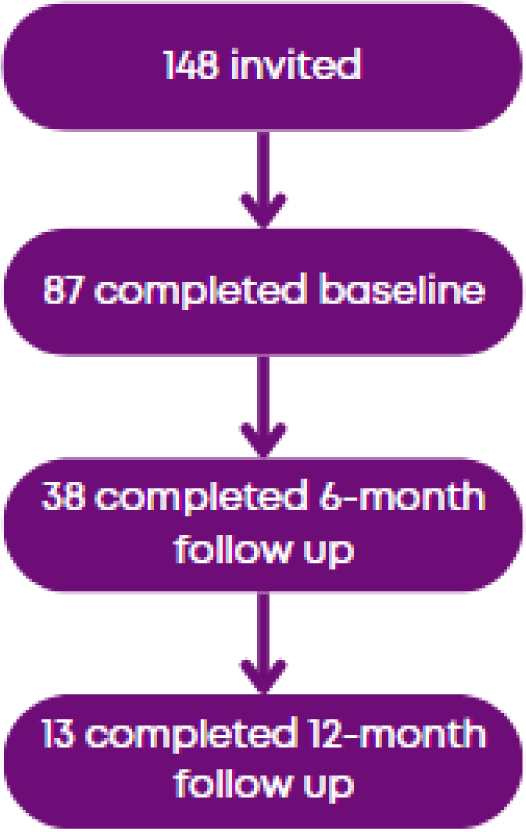
Participation in an evaluation of Sparkle (South Wales)’s specialist leisure provision using the QI-Disability.

## Results

### PedsQL

The response rates for initial completion by eligible children were 26% (N=9) for children and 91% (N=32) for caregivers. Response rates for 3-month follow-up completion were much lower at 3% (N=1) for children/adolescents and 31% (N=10) for caregivers; and response rates for 6-month follow-up completion dropped to 0% for children and 6% (N=2) for caregivers. The baseline total PedsQL mean scores for children were higher than caregiver scores across all three age range versions of the scale. However, it came to light in the evaluation, by an independent observer, that Leisure Support Workers, who were facilitating data collection with the children due to their understanding of the children’s complex needs and communication styles, strongly directed the children’s questions and answers in a bid to help them complete the questionnaire. It was evident that the children had great difficulty understanding the phrasing of the questions, and Leisure Support Workers were thus rephrasing or simplifying questions, which negated the validity of the tool as developed. Thus these results are not reliable, and not reported.

### KINDL^R^

Only 10 children attempted the KINDL^R^ questionnaire, and most were not fully completed. Of those that were complete, some responses may not have been valid as it was unclear if the participants had fully understood the question, or the response that they chose. We had planned to survey many more children, but this attempt was deemed futile in our participant population, as the questions were either too abstract (e.g. ‘I felt on top of the world’ in the 7-13 year old version) or the language was too complex for children with cognitive impairment (e.g., ‘I coped well with the assignments set in nursery’ in the 4-6 year old version). Given these difficulties, we have decided not to present any results from this data collection here.

### QI-Disability

This tool relied exclusively on caregiver responses, with no direct measure for the children to complete. Caregivers were asked to complete the tool prior to the child starting club, and again at 6- and 12-months following commencement of weekly attendance. The results are limited by a significant drop off in respondents at 6 and 12 months, with only 38 (65%) of 6 months scores being completed. As only 13 (21%) caregivers responded to the 12-month questionnaire, this dataset could not be analysed at all. QI-Disability results have been reported in more detail in McGrath et al. (2023). Overall, the only difference found related to ‘positive emotions’ scores, which were lower at baseline (*median* = 75) than after six months of accessing Sparkle leisure services (*median* = 81.25), suggesting an increase in caregiver reports of positive emotions for the children and young people. The large drop off in completion across the different time periods suggested that caregivers found repeat completion of the QI-Disability index onerous, thus potentially limiting its applicability in evaluating a specialist leisure provision.

## Discussion

While the rights of disabled children include their right to play (UNCRC Article 23) (UNICEF, 1992), and there is a clear need for opportunities for children with disabilities to develop social and problem-solving skills, determining the impact of different play opportunities is limited by the lack of appropriate validated tools to measure the impact of such a provision on their QoL. This work has evaluated three well recognised, and previously validated QoL measures (PedsQL, KINDL^R^ and QI-Disability) which are ostensibly suited for a population of children with complex disorders, including cerebral palsy, Autistic Spectrum Disorder, learning difficulties etc. However, it is evident that they are not applicable for children with the most complex needs, and in particular, those with severe communication disorders. In addition, the QI-Disability does not capture the child’s own lived experiences, rather it relies on caregiver responses, thus potentially biasing the results. Although some interesting data was obtained from the QI-Disability, caregivers found it difficult to commit the time to repeat measures, which is unsurprising given the high demands of caring for a child with complex needs. Thus, reliance on caregiver’s completing QoL tools on behalf of their children will inevitably limit the uptake, and value, of these tools.

Quality of life is important to an individual, to their family and to society as a whole, with people who have a higher quality of life experiencing greater happiness and satisfaction in their lives. The importance of play is well documented, with play aiding the development of social skills, language and improving mobility (Alotibi & Algahtani, 2019). Children with complex needs, either physical, cognitive or both, may not be able to experience play in the same way as their unaffected peers and may not be able to access mainstream provision or get the same enjoyment from it. Caregivers responding to the 2018-2019 national survey of children’s health reported that children with special healthcare needs had far lower scores in multiple domains, including making friends and participating in organised activities, than their peers without complex needs (Coleman et al., 2022). This would suggest that there is a need for specialist provision to enable these children and young people to engage and benefit from play activities, to encourage their development, and contribute to their happiness and quality of life. The importance of prioritising wellbeing and quality of life, specifically what matters *most* to children with complex needs, and their families, has been highlighted by the recent ‘A *Blueprint for Change: Guiding Principles for Advancing the System of Services for CYSHCN and their Families*’ (McLellan et al., 2022). However, there is a lack of research on measuring the impact of different forms of leisure provision for this population, or how this may impact their wellbeing or QoL. Often the tools used fail to take account of the child’s cognitive abilities, or communication difficulties, and do not offer them the opportunity to express their own thoughts and feelings. The items being scored also focus heavily on health, rather than overall wellbeing. This evaluation of three well-established tools to assess QoL has shown that existing validated tools are inappropriate to determine the impact of leisure provision for children with complex disability, in particular those with significant communication disorders or cognitive delay.

### Why should we measure quality of life?

Children with intellectual disabilities and autism who have more complex or higher support needs experience poorer quality of life, and *“poorer quality of life can be partly explained by less frequent community participation”* (Williams et al., 2021). There is evidence that suggests the involvement of children with disabilities in groups and clubs can have a positive effect on their skills and emotional wellbeing, in particular their mental health (Brooks et al., 2021). It is more difficult to determine how these groups affect their *overall quality of life*, due in part to the wide range of children and young people’s abilities. Numerous tools have been produced to attempt to measure a child’s quality of life, with or without parent involvement (Hullman et al., 2011). Quality of life and level of disability of a child can have a direct impact on the quality of life of the parent or caregiver. Previous research has shown significant differences between parents with and without a child with disabilities, particularly in social and environmental domains; a positive correlation was shown between increased severity of disability and lower quality of life in the parents (Leung & Li-Tsang, 2003). Improving a child’s quality of life may have a sizeable impact, not only on their wellbeing, but also on their parents, caregivers or families.

However, the current tools available are often aimed at the general population, and fail to take into account children and young people with complex needs, and their capacity for answering the prepared questions, with no provision for those that are non-verbal. Our report assesses previously validated, and widely used, QoL tools in an attempt to evaluate the specialist leisure provision by Sparkle, namely PEDSQL, KINDL^R^ and QI-Disability.

### What is the value of using validated tools to measure QOL in our population?

It is clear from this evaluation that KINDL^R^ and PedsQL were not appropriate tools for use with our population, namely children and young people with significant communication disorders, ASD or cognitive impairment. The wording of the questions was too difficult for our participants to understand, the concepts were too abstract, or they required the children to reflect over the past few weeks in selecting their answer. The PedsQL questions were put to the children by Leisure Support Workers, however an independent observer noted that the questions had to be rephrased/ reworded, and thus the results were likely to be significantly biased due to the level of direction by the support staff. It was not clear with either of these tools whether the children’s responses were meaningful or not. Previous use of this tool has included populations such as those with ADHD, but it is not clear whether these children had significant communication needs or learning difficulties, and the child versions were completed in the home with the parents, and no independent observation took place to determine how much parents may have influenced their child’s responses (Varni & Burwinkle, 2006).

The QI-Disability tool was designed to assess quality of life in children and teenagers who had an intellectual disability, aged between 5 and 18, with design input from the families of affected young people. This tool did not attempt to obtain any information directly from the children, rather it relied on caregiver reporting. Initial conditions included in the derivation of this tool were Down’s and Rett’s syndromes, cerebral palsy (where intellectual disability was present) and autistic spectrum disorder, totalling 77 children and young people involved. As our population of participants primarily had diagnoses of the conditions covered in the initial use of the tool during the design process, it was felt that this would be an appropriate tool to use for evaluation of the services provided. However, it is unclear whether the participants in the initial work had any communication difficulties, and how much input, if any, this group had as individuals in answering the questions, or whether it was solely the caregivers.

The QI-Disability score focuses on six domains derived following a series of primary caregiver interviews (Telethon Kids Institute, 2022), namely general health, positive emotions, negative emotions, social interactions, leisure and the outdoors and independence, with the overall view being quality of life. A limitation of tools which rely solely on caregiver reports to estimate the child’s QoL is that it is well recognised from qualitative research that parents of children with complex needs tend to report lower wellbeing scores than their children (Myers et al., 2021). It was evident within our evaluation that there was a marked drop-off in caregivers returning repeat scoring sheets, which may well indicate that parents of children with complex needs, who spend a disproportionate amount of their time attending appointments and providing personal care to their children throughout the age span, simply do not have the time to complete repeat scores for research purposes. Previous studies have highlighted how ‘time poor’ parents of disabled children are (Leung & Li-Tsang, 2003).

### What’s wrong with the current tools?

Despite having an agreed explanation of what quality of life is, what an individual considers ‘quality’ will vary. What quality looks like for a parent or carer could be considerably different to a child’s perception. Generalised quality domains can be identified, but the significance of a domain to each person may be widely dissimilar. How much do people’s perceptions of what they feel constitutes a good quality of life are projected onto their assessment of another? Children with cerebral palsy and their parents highlighted a number of areas that were important for a good quality of life, for example physical activity, but when you consider the differences in ability between a child with cerebral palsy Gross Motor Function Classification System 1 (GMFCS) and another with GMFCS 4 or 5, it is highly likely that what they would consider quality of activity would be different (Davies et al., 2017).

Previous studies, e.g. using the KINDL questionnaire, have shown significant differences between a child’s score and a mother’s opinion score (Rotsika et al., 2011), therefore demonstrating that parents may not be able to give a correct reflection of their child’s quality of life, although this study compared those with ‘specific learning difficulties’ to ‘typically developing’ children, they all had a normal IQ, thus do not reflect those with severe complex disability. Likewise, Klassen et al. (2006) reported a higher QoL for children with Attention Deficit Hyperactivity Disorder, than that scored by their caregivers, using the Child Health Questionnaire (CHQ). There was agreement between children and caregivers in this study relating to physical health rather than psychosocial health. We wish to identify a QoL tool that explicitly explores aspects of the child’s emotional wellbeing and happiness, rather than their physical limitations, which are far less likely to be altered by attendance at a specialist leisure provision. The CHQ was discounted as a potential tool for our families, as the child version (age 10-18 years) has 87 items, and the validation has not been conducted explicitly on those with learning difficulties or complex needs. It is not clear whether the caregiver is able to truly estimate their child’s QoL, as what may be of value to the child, may not seem valuable to the caregiver. Furthermore, given the considerable pressure that parents of children with complex needs are under, it is possible that their scoring may in fact reflect their own mood/ QoL, rather than that of their child. Of note, parents of children with cerebral palsy report lower quality of life scores than the children themselves (Makris et al., 2021). Previous published research from Sparkle looked at the impact of residential trips on adolescents with complex needs and in this instance the young people consistently scored themselves higher for enjoyment and improvement in social skills than their parents and caregivers (Myers et al., 2021). Both these examples highlight the need for an accessible measurement scale. Makris et al. (2021) recommends multi-source measures including both the child’s and parent/carer’s views, as well as the views of professionals working with the family, as a gold standard approach to measuring QoL, which we have been exploring as a research method for our group (Collins et al., 2023). It is vital when assessing the value or impact of any provision to obtain the views of *those actually using and experiencing the service*, particularly those with less self-agency, whose views are rarely sought (Cheak-Zamora et al., 2015). One of the difficulties of measuring the user’s quality of life in this population is the child’s ability to communicate their thoughts and feelings in a manner that can be standardised and represented. There are numerous strategies in place for children and young people to communicate, i.e., Makaton or Picture Exchange Communication System (PECS), all of which are used by the Sparkle leisure team, but these do not lend themselves to answering the often complex questions laid out in quality of life evaluation tools. Future research developing standardised tools for this population need to consider a greater simplification of required answers to ensure they are applicable to children with more cognitive delay, or rigid thinking.

### Limitations

While usable data were obtained using the QI-Disability tool, this was not without limitations. Only 13 of the initial caregivers completed the 12-month follow-up (22%), whereas the 6-month follow-up was completed by 65%, limiting the statistical analyses that could be conducted. While the trend at 12 months appeared to follow the initial trajectory, its validity was limited by a much smaller participant population than originally planned.

An additional potential complication was that our work using the QI-Disability tool began in the aftermath of the COVID-19 pandemic. During the pandemic, the specialist leisure clubs did not run for six months, and many of the children that usually participated in the clubs would have been classed as vulnerable. It is possible to hypothesise that due to this group’s susceptibility to COVID-19, not only were they not accessing or had delayed access to the leisure clubs, but that they found themselves more isolated than their mainstream peers. Research during and after the pandemic, and subsequent lockdowns, suggested over 90% of children and young people were socially isolated with over half reported by their parents to have had a negative impact on social skills and over 70% experiencing more frequent negative emotions (Lunt, 2021). Loneliness and isolation was a common feature, with 72% of adults with intellectual impairment feeling anxious or down, and 41% feeling lonely with no one to talk to (Flynn et al., 2021). If the children and young people were isolated from others within their usual activities and educational settings, it may be that starting at Sparkle leisure activities they had a lower baseline across the domains than they may have done without pandemic isolation. It could therefore be suggested that an even longer length of time may needed to see improvements in complex skills like socialisation, even accounting for the difficulties that some might have with this due to their underlying diagnosis e.g. Autistic Spectrum Disorder.

### Implications

Every child and young person has a right to accessible play activities, to not only boost their physical health but also their mental wellbeing and improve skills acquisition. The WHO has recently updated guidance on the benefits of physical activity for young people, with direct mention of those with disabilities being able to improve their physical health, cognitive functioning and mental health (WHO, 2022). Specialist provision provides an appropriate setting for activity and play for this group of children, but comes at a higher cost. Children with complex needs often require more specialist equipment and more specialist staff support than mainstream activities, and at a time of increasing cost of living it is vital that any services provided are of a high standard, with a demonstrable impact on the children, and their wider family, not only to ensure appropriate use of funds, but also to provide the best available service for their users.

Being able to measure the impact of such provision is vital to demonstrating the high standards needed and expected, and the impact of the provision. Parents and carers of children with additional needs already have significant demands on their time, so any tools need to be quick and easy to use with results displayed in an easy to access and comprehensible format to ensure the exercise is worthwhile, and representative of the population. As previously discussed, discrepancies may exist between parent-reported and child-reported measures, thus there is a need for future research to gather and understand the experience of the children and young people as service users, and to explore other ways of evaluating services and the needs of their users.

### How should we measure quality of life in this population?

The United Nations Convention on the Rights of a Child states that a child has the right to give an opinion on important decisions that affect them, the right to find out information and the right to think for themselves. Despite this, there is a lack of instruments and mechanisms for a child with a cognitive or communicative disability to have their voice heard. Children and young people with disabilities and their families are best placed to explain and explore what they need and want from specialist leisure provision and indeed the health service (Coleman et al., 2022). Quality of life and health should factor in so much more than disability and encompass other attributes like social skills and play. Developing appropriate tools to meet this requires systemic change and recognition of their importance in the lives of those affected.

### Play is a right for all children, as is having their voice heard

Increasing rates of childhood disability will place increasing demands on service providers to meet the needs of this group (Dalgaard et al., 2022). In order to determine if this specialist provision is appropriate, and having the desired impact on children’s wellbeing and development, appropriate tools are required.

This work was conducted as part of a real-world service, and not a randomised controlled trial, and therefore there are challenges to the evaluation of such services. Despite the many advancements in providing specialist care for children and young people with physical and cognitive disabilities, there still remains a gap for an accessible and inclusive quality of life evaluation tool. Until it’s development we continue to rely on those that know the child or young person best and the identification of non-verbal cues. More needs to be done to include this population of children and young people in research and allow their opinions to matter and be heard.

## Data availability

Data are not openly available due to lack of participant consent to open the data.

## About the authors

Dr Fiona Astill is a ST6 Community Child Health Registrar at Aneurin Bevan University Health Board, based at St Cadoc’s Hospital. Bethan Collins MSc is the Research and Development Officer at Sparkle (South Wales), based at Serennu Children’s Centre in Newport, South Wales. Dr Nicole McGrath is a Consultant Paediatrician at Aneurin Bevan University Health Board, based at Serennu Children’s Centre. Professor Alison Kemp is an Emeritus Professor at Cardiff University School of Medicine. Dr Lisa Hurt is a Senior Lecturer in maternal and child health epidemiology at Cardiff University School of Medicine, Division of Population Medicine. Dr Sabine Maguire is an Honorary Research Fellow at Cardiff University, and an unsalaried trustee of Sparkle (South Wales).

## Acknowledgements

We would like to thank the children, caregivers and leisure support workers who assisted in evaluating these tools. We are also grateful to Chloe Hurrell for her independent evaluation of the PedsQL tool.

## Plain English Summary

Children and young people (CYP) with complex needs deserve to have the opportunity to experience a quality of life (QoL) equal to that of their non-disabled peers. Measuring QoL in this group is problematic and there is yet to be developed an appropriate tool for allowing their opinions to be heard. Many existing tools use parents or carers to answer for their CYP, which may often be different to how they would score themselves, and carers have many demands on their time, thus are less likely to participate in repeated questionnaires. This study assessed three different previously validated tools to measure QoL in our population of children and young people accessing specialist leisure provision, and found them wanting. There is a paucity of validated tools for children with significant communication difficulties and/ or cognitive impairment, which means that their voice is not being heard. More research is needed to explore the quality of life that these young people are experiencing, and whether the provision they receive is benefitting them.

## Appendix 1 Aims of the specialist leisure clubs delivered by Sparkle (South Wales)

**Appendix 1 - Aims of the specialist leisure clubs delivered by Sparkle (South Wales).**
| *Club* | *Aims of clubs* |
| --- | --- |
| Youth Clubs | Independence, social skills, communication skills, interacting with others/ building relationships |
| Play Clubs | Play opportunities, children engaging with resources, social skills, communication, friendships |
| After School Club (5-11 years) | Play opportunities, children engaging with resources, social skills, communication, friendships |
| After School Club (12-17 years) | Independence, social skills, communication skills, interacting with others/ building relationships |
| Skills Club | Physically active, building stamina, co-ordination and social skills, team work |
| Independent Living Skills | Independence, skills for independent living |
| Technology Club | Develop skills in technology, interaction with peers, communication skills, turn taking, managing frustrations (gaming related) |

## References

Alotibi, A. & Algahtani, F. (2019). The impact of play on quality of life on children with intellectual disability. International Journal of Recent Research in Social Sciences and Humanities, 6(4), 84–91, https://www.paperpublications.org/upload/book/The%20impact%20of%20play-1442.pdf

Brooks, R., Lambert, C., Coulthard, L., Pennington, L. & Kolehmainen, N. (2021). Social participation to support good mental health in neurodisability. Child: Care, Health and Development, 47(5), 675–684, 10.1111/cch.12876

Cheak-Zamora, N. C., Teti, M. & First, J. (2015). ‘Transitions are Scary for our Kids, and They’re Scary for us’: Family Member and Youth Perspectives on the Challenges of Transitioning to Adulthood with Autism. Journal of Applied Research in Intellectual Disabilities, 28(6), 548–560, 10.1111/jar.12150

Coleman, C. L., Morrison, M., Perkins, S. K., Brosco, J. P. & Schor, E. L. (2022). Quality of Life and Well-Being for Children and Youth With Special Health Care Needs and their Families: A Vision for the Future. Pediatrics, 149(7), 10.1542/peds.2021-056150G

Collins, B., McGrath, N., Astill, F., Hurt, L., Maguire, S. & Kemp, A. (2023). Meaningful outcomes of specialist leisure activities for children with complex disabilities: the views of parents, professionals and young people. MedRxiv, 10.1101/2023.11.15.23298514

Dahan-Oliel, N., Shikako-Thomas, K. & Majnemer, A. (2012). Quality of life and leisure participation in children with neurodevelopmental disabilities: a thematic analysis of the literature. Quality of Life Research, 21(3), 427–439, 10.1007/s11136-011-0063-9

Dalgaard, N.T., Bondebjerg, A., Viinholt, B. C. A. & Filges, T. (2022). The effects of inclusion on academic achievement, socioemotional development and the wellbeing of children with special educational needs. Campbell Systematic Reviews, 18(4), 10.1002/cl2.1291

Davis, E., Reddihough, D., Murphy, N., Epstein, A., Reid, S. M., Whitehouse, A., Williams, K., Leonard, H. & Downs, J. (2017). Exploring quality of life of children with cerebral palsy and intellectual disability: What are the important domains of life? Child: Care, Health and Development, 43(6), 854–860, 10.1111/cch.12501

Downs, J., Jacoby, P., Leonard, H., Epstein, A., Murphy, N., Davis, E., Reddihough, D., Whitehouse, A. & Williams K. (2019). Psychometric properties of the Quality of Life Inventory-Disability (QI-Disability) measure. Quality of Life Research, 28(3), 783–794, 10.1007/s11136-018-2057-3

Flynn, S., Hayden, N., Clarke, L., Caton, S., Hatton, C., Hastings, R. P., Abbott, D., Beyer, S., Bradshaw, J., Gillooly, A., Gore, N., Heslop, P., Jahoda, A., Maguire, R., Marriott, A., Oloidi, E., Paris, A., Mulhall, P., Scior, K., Taggart, L., & Todd, S. (2021). Coronavirus and People with Learning Disabilities Study: Wave 3 Results. Retrieved from https://pure.ulster.ac.uk/ws/portalfiles/portal/92511950/Coronavirus_and_People_with_Learning_Disabilities_Study_Wave_3_Full_Report_v1.0_FINAL.pdf

Graham, N., Nye, C., Mandy, A., Clarke, C. & Morriss-Roberts, C. (2018). The meaning of play for children and young people with physical disabilities: A systematic thematic synthesis. Child: Care, Health and Development, 44(2), 173–182, 10.1111/cch.12509

Hullman, S. E., Ryan, J. L., Ramsey, R. R., Chaney, J. M. & Mullins, L. L. (2011). Measure of general pediatric quality of life: Child Health Questionnaire (CHQ), DISABKIDS Chronic Generic Measure (DCGM), KINDL-R, Pediatric Quality of Life Inventory (PedsQL), 4.0 Generic Core Scales, and Quality of My Life Questionnaire (QoML). Arthritis Care and Research, 63(11), 420–430, 10.1002/acr.20637

Klassen, A. F., Miller, A. & Fine, S. (2006). Agreement between parent and child report of quality of life in children with attention-deficit/hyperactivity disorder. Child: Care, Health and Development, 32(4), 397–406, 10.1111/j.1365-2214.2006.00609.x

Kramer, S., Moller, J. & Zimmermann, F. Z. (2021). Inclusive Education of Students with General Learning Difficulties: A Meta-Analysis. Review of Educational Research, 91(3), 432–478, 10.3102/0034654321998072

Leung, C. & Li-Tsang, C. (2003). Quality of Life of Parents who have Children with Disabilities. Hong Kong Journal of Occupational Therapy, 13(1), 19–24, 10.1016/S1569-1861(09)70019-1

Lewis, J. M. (1993). Childhood play in normality, pathology, and therapy. American Journal of Orthopsychiatry, 63(1), 6–15, 10.1037/h0079403

Longo, E., Badia, M., Begona-Orgaz, M. & Gomez-Vela, M. (2017). Comparing parent and child reports of health-related quality of life and their relationship with leisure participation in children and adolescents with Cerebral Palsy. Research in Developmental Disabilities, 71, 214–222, 10.1016/j.ridd.2017.09.020

Lunt, C. (2021). The Loneliest Lockdown – The Impact of the Pandemic on Disabled Children, their Parents and Siblings. Disabled Children’s Partnership. Retrieved from https://disabledchildrenspartnership.org.uk/wp-content/uploads/2021/03/The-Loneliest-Lockdown.pdf

Makris, T., Dorstyn, D. & Crettenden, A. (2021). Quality of life in children and adolescents with cerebral palsy: a systematic review with meta-analysis. Disability and Rehabilitation, 43(3), 299–308, 10.1080/09638288.2019.1623852

McGrath, N., Astill, F., Collins, B., Maguire, S., Kemp, A. & Hurt, L. (2023). The effect of leisure activities on quality-of-life scores for children with complex needs: A service evaluation in Wales, UK. MedRxiv, 10.1101/2023.12.22.23300435

McLellan, S. E., Mann, M. Y., Scott, J. A. & Brown, T. W. (2022). A Blueprint for Change: Guiding Principles for a System of Services for Children and Youth With Special Health Care Needs and Their Families. Pediatrics, 149(7), 10.1542/peds.2021-056150C

Myers, S., Elliot, F., Maguire, S. & Barber, M. (2021). Improving Social-Emotional and Life Skills of Young People with Complex Additional Needs Through ‘Outward Bound’ Residential Trips. International Journal of Disability, Development and Education, 70(6), 10.1080/1034912X.2021.1944992

Ravens-Sieberer, U. & Bullinger, M. (1998). Assessing health-related quality of life in chronically ill children with the German KINDL: First psychometric and content analytical results. Quality of Life Research, 7(5), 399–407, 10.1023/a:1008853819715

Rotsika, V., Coccossis, M., Vlassopoulos, M., Papaeleftherious, E., Sakellariou, K., Anagnostopoulos, D. C., Kokkevi, A. & Skevington, S. (2011). Does the subjective quality of life of children with specific learning disabilities (SpLD) agree with their parents’ proxy reports? Quality of Life Research, 20(8), 1271–1278, 10.1007/s11136-011-9857-z

Shikako-Thomas, K., Dahan-Oliel, N., Shevell, M., Law, M., Birnbaum, R., Rosenbaum, P., Poulin, C. & Majnemer, A. (2012). Play and be happy? Leisure participation and quality of life in school-aged children with cerebral palsy. International Journal of Paediatrics, 10.1155/2012/387280

Telethon Kids Institute. QI-Disability. Accessed 17th October 2022. Available from: https://www.telethonkids.org.au/our-research/brain-and-behaviour/disability/child-disability/qi-disability/

UNICEF. (1992). A Summary of the UN Convention on the Rights of the Child. Retrieved from https://www.unicef.org.uk/wp-content/uploads/2019/10/UNCRC_summary-1_1.pdf

Varni, J. W. & Burwinkle, T. M. (2006). The PedsQLTM as a patient-reported outcome in children and adolescents with attention-deficit/hyperactivity disorder: A population-based study. Health and Quality of Life Outcomes, 4, 26, 10.1186/1477-7525-4-26

Whitebread, D. (2012). The Importance of Play. Retrieved from https://www.csap.cam.ac.uk/media/uploads/files/1/david-whitebread---importance-of-play-report.pdf

Williams, K., Jacoby, P., Whitehouse, A., Kim, R., Epstein, A., Murphy, N., Reid, S., Leonard, H., Reddihough, D. & Downs, J. (2021). Functioning, participation, and quality of life in children with intellectual disability: an observational study. Developmental Medicine and Child Neurology, 63(1), 89–96, 10.1111/dmcn.14657

World Health Organisation [WHO]. (2022). Physical activity. Retrieved from https://www.who.int/news-room/fact-sheets/detail/physical-activity#:∼:text=Children%20and%20adolescents%20aged%205,physical%20activity%2C%20across%20the%20week.

World Health Organisation [WHO]. (2022). WHOQOL: Measuring Quality of Life. Retrieved from https://www.who.int/tools/whoqol

